# Incidence of and Risk Factors for New-Onset Urinary and Fecal Incontinence After Acute Stroke

**DOI:** 10.1101/2023.09.11.23295391

**Authors:** Enrique Cruz, Yvonne Wells, Charne Miller, Natasha A. Lannin, Geoffrey C. Cloud

**Author notes:** CORRESPONDING AUTHOR: Enrique Cruz; Address: Caulfield Hospital, 260 Kooyong Road, Caulfield VIC Australia 3162.

## Abstract

**Background and Purpose:** Understanding the risk factors for incontinence after acute stroke allows for early identification and treatment of patients who are likely to experience this complication. This study estimates incidence of urinary and fecal incontinence (UI and FI) after stroke in adults and examines the association of risk factors.

**Methods:** Screening data of a prospective study were analyzed. Included were 652 consecutive patients admitted to an acute stroke unit over a 15-month period beginning May 2021. Those with pre-existing UI or FI were excluded from the analysis. Bivariate analyses examined unadjusted relationships between both UI and FI and age, sex, stroke severity, premorbid disability, stroke type, and stroke anatomical site. Regression analyses were performed to build iterative models.

**Results:** A total of 418 patients had an acute stroke without pre-existing incontinence. The median age of stroke survivors was 76 years (*IQR* 66-84), 58% were men, and 76% had an ischemic stroke. Within the first 7 days after stroke, 128 (29%) had UI and 115 (26%) had FI. Increasing age, premorbid disability, and stroke severity increased the risk of both new-onset UI and FI after stroke. Those with intraventricular hemorrhage were 2.9 times more likely to develop UI. Women were 1.7 times more likely to have UI after stroke than men. Intraventricular hemorrhage increased the risk of new-onset FI after stroke by 2.9 times, while lobar hemorrhage increased the risk by 2.2 times.

**Conclusions:** Urinary and fecal incontinence continue to be relatively common after acute stroke, although incidence rates are lower than 20th century estimates. Patients are more likely to require incontinence assessment, monitoring and care if they are older, had higher premorbid disability, and have experienced more severe stroke or intraventricular hemorrhage. Provision of continence care and rehabilitation remain essential components of stroke services.

## INTRODUCTION

Incontinence, defined as involuntary leakage of urine or feces, is a stroke complication that reduces a patient’s ability to be independent in personal care, enjoy leisure activities, and interact with others^1^. Stroke patients with incontinence resume fewer of their pre-stroke activities than those without^2^. This decline in function can be attributed to the physical and psychological difficulties that incontinence presents. Increased severity of incontinence after stroke is associated with decreased quality of life^3^ and increased dependence on carers. The need for assistance can lead to feelings of embarrassment and loss of dignity^4^.

Knowing the risk factors for stroke complications such as incontinence enables early identification and prompt treatment^5^, including proactive measures to assess and manage continence issues. Early identification allows for timely interventions such as medication review, pelvic floor exercises, and bladder training^6^. Prompt treatment can significantly improve the quality of life for stroke survivors and reduce the burden of incontinence on both patients and caregivers^7^. Understanding the risk factors for, and incidence of, incontinence after stroke enables focused screening and surveillance of at-risk groups and guides resource allocation.

Community-based studies about the incidence of, and risk factors for, incontinence after stroke are few and lack currency. The effectiveness of acute stroke treatments has improved considerably in the last decade, and the incidence of stroke complications such as incontinence may have changed. Between 1990 to 2020, estimates of the incidence and prevalence of urinary incontinence (UI) after stroke were 44.3%^8^ and 43.5 to 47%^9–11^ respectively. In the same period, the estimates of incidence and prevalence of fecal incontinence (FI) after stroke were 30%^12^ and 40%^11^ respectively. Results of a recent New Zealand study suggest the incidence of UI and FI among adults presenting to hospital after stroke may have decreased^13^.

The aim of this analysis was to provide updated incidence estimates of UI and FI among adults admitted to acute care after stroke and to examine the association of previously published risk factors for new-onset UI and FI after acute stroke.

## METHODS

We analyzed the screening data of a single-center prospective intervention study on incontinence after stroke that was registered with the Australian New Zealand Clinical Trials Registry (ANZCTR; Registration number ACTRN12621000879864).

This study was approved by the ethics committee of Alfred Health in Melbourne, Australia (Reference number 57/20). Included were 652 inpatients admitted consecutively to the acute stroke unit of a Comprehensive Stroke Centre (CSC) during a 15-month period beginning 1 May 2021. The patients were followed-up until they were discharged from the hospital.

Stroke was diagnosed by a consultant stroke physician according to the International Classification of Diseases 11^th^ revision (ICD-11) criteria; diagnosis required the presence of acute neurological dysfunction. Stroke types included ischemic, intracerebral hemorrhage, subarachnoid hemorrhage, and stroke not known if ischemic or hemorrhagic based on brain imaging reports. Computed Tomography (CT) scans were performed using a Canon Aquilion scanner (Canon Medical System, USA), and follow-up Magnetic Resonance Imaging (MRI) was performed using a Siemens Magnetom scanner (Siemens Healthineers, Germany). The type and anatomical location of the stroke were determined based on reports provided by a consultant radiology physician and subsequently verified by a consultant stroke physician.

Incontinence was evaluated by a continence nurse using International Continence Society (ICS) definitions^14^. Urinary incontinence was defined as the involuntary loss of urine and fecal incontinence was the involuntary loss of feces. History of incontinence was also assessed. Those who experienced incontinence four weeks prior to their stroke were classified as having pre-morbid incontinence.

Stata software (StataCorp, USA) was used for statistical analysis. Bivariate analyses explored unadjusted associations between UI and FI and age, sex, stroke severity (using the NIHSS), premorbid disability (using the mRS), type of stroke, and anatomic location of stroke. Variables that were statistically significant predictors of new-onset incontinence (*P* < 0.10) were included in logistic and backwards stepwise regression analyses (*P* to enter = 10%, *P* to remove = 5%) to build iterative models. Wald tests were used to assess potential interactions between covariates, and Hosmer-Lemeshow tests were performed to assess goodness-of-fit.

## RESULTS

During the study period, 652 individuals were admitted to the acute stroke unit, 440 of whom had a confirmed diagnosis of acute stroke. The median age of stroke survivors was 76 years (*IQR* 66-84); 58% were men, and 76% had experienced ischaemic stroke. The median National Institutes of Health Stroke Scale (NIHSS) score was 4 (*IQR* 2-12), and median modified Rankin Scale (mRS) score was 0 (*IQR* 0-1). The demographic and clinical characteristics of the acute stroke patients are shown in Table 1.

**Table 1:**
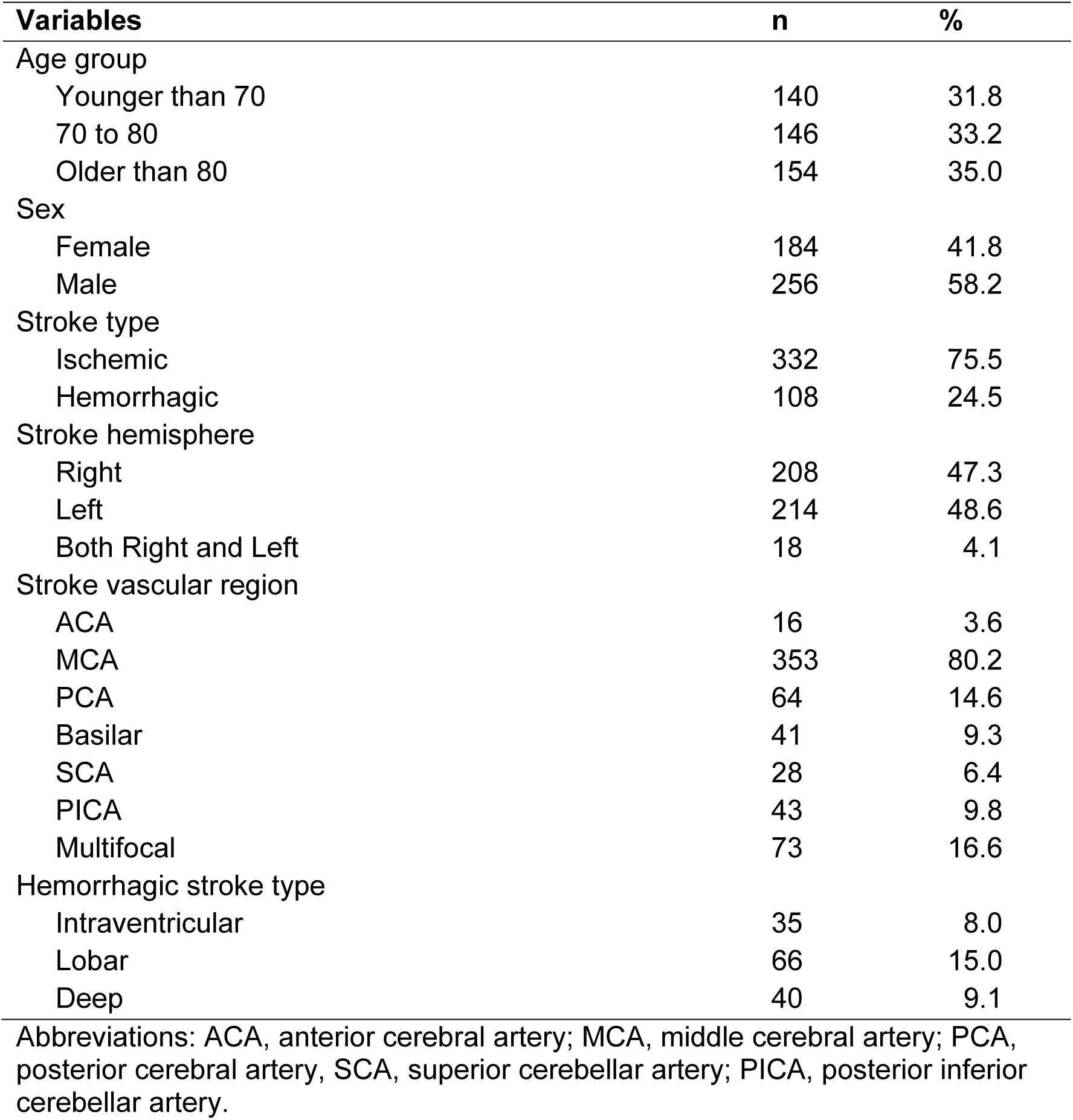
Characteristics of Patients with Acute Stroke (n = 440)

| Variables | n | % |
| --- | --- | --- |
| Age group |  |  |
| Younger than 70 | 140 | 31.8 |
| 70 to 80 | 146 | 33.2 |
| Older than 80 | 154 | 35.0 |
| Sex |  |  |
| Female | 184 | 41.8 |
| Male | 256 | 58.2 |
| Stroke type |  |  |
| Ischemic | 332 | 75.5 |
| Hemorrhagic | 108 | 24.5 |
| Stroke hemisphere |  |  |
| Right | 208 | 47.3 |
| Left | 214 | 48.6 |
| Both Right and Left | 18 | 4.1 |
| Stroke vascular region |  |  |
| ACA | 16 | 3.6 |
| MCA | 353 | 80.2 |
| PCA | 64 | 14.6 |
| Basilar | 41 | 9.3 |
| SCA | 28 | 6.4 |
| PICA | 43 | 9.8 |
| Multifocal | 73 | 16.6 |
| Hemorrhagic stroke type |  |  |
| Intraventricular | 35 | 8.0 |
| Lobar | 66 | 15.0 |
| Deep | 40 | 9.1 |

Abbreviations: ACA, anterior cerebral artery; MCA, middle cerebral artery; PCA, posterior cerebral artery; SCA, superior cerebellar artery; PICA, posterior inferior cerebellar artery.

Among those who had acute stroke, 22 (5%) had premorbid incontinence and were excluded from the analysis (see Figure 1,CONSORT flow diagram). The median length of stay in the acute stroke unit is 6 days (*IQR* 3-10).

**Figure 1:**
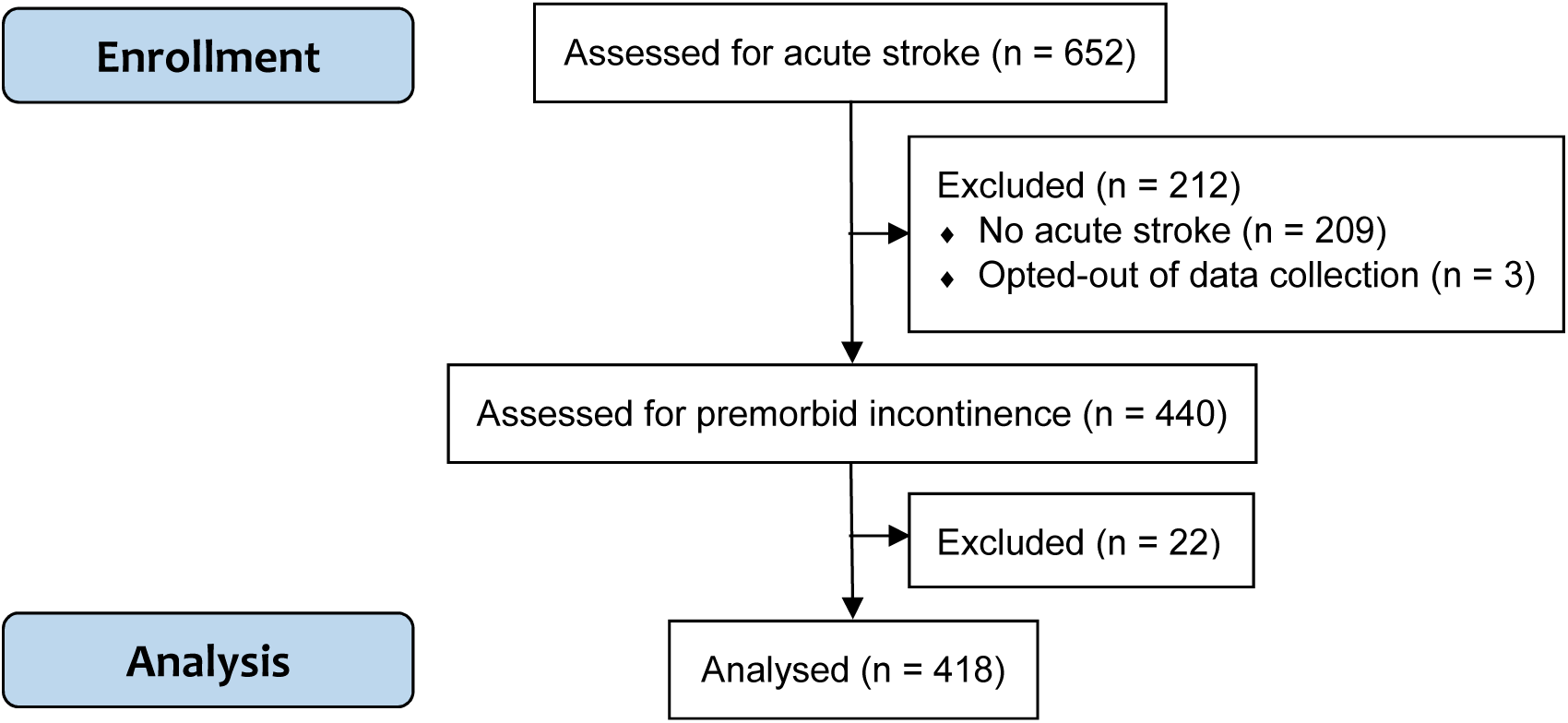
CONSORT Flow Diagram.

Within seven days of stroke, 107 (26%) developed new-onset UI and 103 (25%) developed new-onset FI. Results of bivariate analysis of the characteristics of patients with and without new-onset incontinence are shown in Table 2. There was a notable difference in the discharge outcomes between those who developed incontinence after acute stroke and those who did not, with most patients without incontinence discharged back to their homes (see Table 3).

**Table 2:** Bivariate Logistic Regression Analysis of Unadjusted Association Between Patient Characteristics and Incidence of Incontinence After Stroke.

| Characteristics | New-onset UI |  |  | New-onset FI |  |  |
| --- | --- | --- | --- | --- | --- | --- |
|  | OR | 95% CI | P | OR | 95% CI | P |
| Age (years) | 1.06* | 1.03-1.08 | <0.01 | 1.05* | 1.03-1.08 | <0.01 |
| Sex - Female | 2.02* | 1.29-3.15 | <0.01 | 1.55 | 0.99-2.42 | 0.55 |
| mRS | 1.76* | 1.48-2.10 | <0.01 | 1.80* | 1.50-2.15 | <0.01 |
| NIHSS | 1.20* | 1.16-1.25 | <0.01 | 1.20* | 1.15-1.24 | <0.01 |
| Hemorrhagic stroke | 2.21* | 1.37-3.58 | <0.01 | 2.70* | 1.67-4.37 | <0.01 |
| Stroke hemisphere |  |  |  |  |  |  |
| Right | 1.25 | 0.81-1.95 | 0.32 | 1.08 | 0.69-1.69 | 0.73 |
| Left | 0.85 | 0.55-1.31 | 0.46 | 0.99 | 0.63-1.54 | 0.96 |
| Vascular region |  |  |  |  |  |  |
| ACA | 1.79 | 0.63-5.04 | 0.27 | 1.89 | 0.67-5.32 | 0.23 |
| MCA | 2.34* | 1.22-4.51 | 0.01 | 2.21* | 1.14-4.26 | 0.02 |
| PCA | 0.74 | 0.38-1.42 | 0.37 | 0.62 | 0.31-1.24 | 0.18 |
| Basilar | 0.68 | 0.30-1.52 | 0.35 | 0.49 | 0.20-1.21 | 0.12 |
| SCA | 0.97 | 0.39-2.34 | 0.94 | 0.82 | 0.32-2.09 | 0.68 |
| PICA | 0.47 | 0.19-1.15 | 0.10 | 0.49 | 0.20-1.21 | 0.12 |
| Multifocal | 0.94 | 0.52-1.69 | 0.84 | 0.83 | 0.45-1.53 | 0.55 |
| Hemorrhage subtype |  |  |  |  |  |  |
| Intraventricular | 6.27* | 2.89-13.59 | <0.01 | 7.81* | 3.54-17.24 | <0.01 |
| Lobar | 1.86* | 1.05-3.28 | 0.03 | 2.54* | 1.45-4.45 | <0.01 |
| Deep | 2.31* | 1.16-4.59 | 0.02 | 2.16* | 1.08-4.33 | 0.03 |

Abbreviations: UI, urinary incontinence; FI fecal incontinence; OR, odds ratio; CI, confidence interval; mRS, modified rankin scale; NIHSS, national institutes of health stroke scale; ACA, anterior cerebral artery; MCA, middle cerebral artery; PCA, posterior cerebral artery; SCA, superior cerebellar artery; PICA, posterior inferior cerebellar artery.
\*Statistically significant

**Table 3:** Discharge Outcomes of Patients with Acute Stroke Stratified by Incidence.

| **Outcome** | **New-onset UI** |  | **New-onset FI** |  |
| --- | --- | --- | --- | --- |
|  | **With**<br>**(n=107)** | **Without**<br>**(n = 311)** | **With**<br>**(n=103)** | **Without**<br>**(n = 315)** |
| Died in hospital or discharged with end-of-life care service | 41% | 6% | 46% | 5% |
| Discharged to residential care service facility | 35% | 9% | 31% | 11% |
| Discharged home with or without support services | 24% | 85% | 23% | 84% |

Abbreviations: UI, urinary incontinence; FI, fecal incontinence.

Statistically significant variables (*P* ≤ 0.10) were included in multivariate logistic regression analysis, following backward stepwise selection (*P* to enter = 10%, *P* to remove = 5%) to identify risk factors for new-onset incontinence. Separate models were fitted for urinary and fecal incontinence, and the significant risk factors are presented in Tables 4 and 5 respectively.

**Table 4:** Multivariate Logistic Regression Model of Predictors of New-Onset Urinary.

| **Factors** | **OR** | **95% CI** | **P** |
| --- | --- | --- | --- |
| Age | 1.03 | 1.00 - 1.05 | 0.02 |
| Sex - Female | 1.75 | 1.01 - 3.02 | 0.04 |
| mRS | 1.33 | 1.07 - 1.65 | 0.01 |
| NIHSS | 1.17 | 1.12 - 1.22 | <0.01 |
| Intraventricular hemorrhage | 2.89 | 1.13 - 7.40 | 0.03 |

Abbreviations: OR, odds ratio; CI, confidence interval; mRS, modified rankin scale; NIHSS, national institutes of health stroke scale.
\**n* = 418; *R-squared*: 0.2959.

**Table 5:**
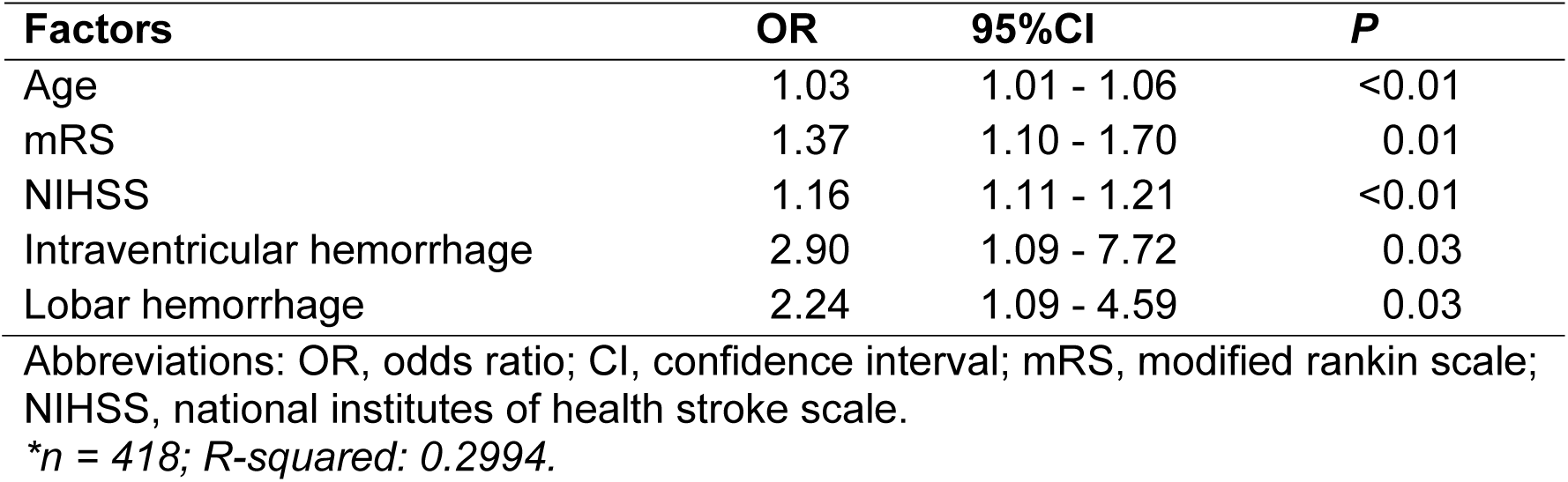
Multivariate Logistic Regression Model of Predictors of New-Onset Fecal.

In the model, those with intraventricular hemorrhage were 2.9 times more likely to develop new-onset UI than those who did not have intraventricular hemorrhage after stroke. Women were 1.7 times more likely to have new-onset UI after stroke than men. Increasing age, premorbid disability, and stroke severity also increased the risk of new-onset UI after stroke.

Intraventricular hemorrhage also resulted in a 2.9-fold increase in the risk of new-onset FI after stroke. Those with lobar hemorrhage were 2.2 times more likely to develop new-onset FI after stroke than those without lobar hemorrhage. As with new-onset UI, increasing age, premorbid disability, and stroke severity also increased the risk of new-onset FI after stroke.

## DISCUSSION

The incidence of new-onset UI after stroke found in this study was lower than previously published estimates (i.e., 26% vs. 44%^8^). Our finding is consistent with a 2022 New Zealand study that found lower-than-expected rates of UI after stroke^13^. Similarly, the incidence of new-onset FI after stroke found in this study (25%) was marginally lower than published estimates (30%^12^). Potentially of importance is that our study site was an acute stroke unit that provide organized, multidisciplinary stroke services. There is compelling evidence that such stroke units can prevent stroke complications^15^, which may have led to lower incident rates.

The prevalence of UI preceding stroke observed in our study (4.8%) was similar to the prevalence observed in England; 7.3% in 2018^10^ and 2.5% in 1989^16^. However, our estimate (4.8%) was much lower than 2012 Australian^9^ and 1986 New Zealand estimates^17^ (14.3% and 17% respectively). Differences in research methods may contribute to explaining these variations, as the 2012 Australian study included only first-ever strokes^9^, and the 1986 New Zealand study^17^ utilized a definition of stroke that differed from the ICD-11 criteria.

The prevalence of any UI after acute stroke (premorbid and new-onset) found in this study (29%) was lower than published estimates (44 to 47%^9–11^). The prevalence of any FI after stroke (premorbid and new-onset) was also lower than in published data (26% vs. 40%^11^). The prevalence of any UI after stroke in Australasian settings appears to have declined over time, from 60% in 1986^17^, to 43.5% in 2012^9^, to 29% in our study. Similarly, the prevalence of any FI after stroke appears to have declined with time, from 40% in 1997^11^, to 30% in 2003^12^, to 26% in our study.

Data on urinary and fecal incontinence, such as those obtained in this study, are typically accessible only through audits^18^ and are yet to be routinely collected in national stroke registries such as the Australian Stroke Clinical Registry (AuSCR)^19^. It is recommended that pre-morbid and post-stroke urinary UI and FI status be included in national clinical and quality care stroke registries to maintain the usefulness of these incidence and prevalence estimates in planning for stroke services and research.

The majority of the current study’s findings on the risk factors for UI after a stroke are consistent with those identified by previous research^8,11,20,21^, including increasing stroke severity, advancing age, increasing pre-morbid disability, and having experienced hemorrhagic stroke.

In our study, female sex was found to be a significant risk factor for new-onset urinary incontinence after stroke (see table 4); however, this was not observed in a previous study^20^. Stroke anatomical location did not significantly predict new-onset incontinence after stroke in this study; however, previous studies have suggested those with lacunar infarcts are less likely to have UI after stroke^20^ and those with stroke lesions in the parietal lobe are more likely to develop UI after stroke^8^.

Further, our multivariate risk factor analysis of new-onset UI and FI after stroke identified intraventricular hemorrhage as a potential risk factor for UI and FI after stroke. This new finding resembles those that found normal pressure hydrocephalus is associated with both urinary^22^ and fecal^23^ incontinence. Overall, the risk factors identified in the current study are consistent with the major mechanisms of UI after stroke^24,25^, which include: (1) direct injury to the neuromicturition pathways, (2) detrusor hyporeflexia with overflow incontinence, (3) impaired cognition UI, (4) impaired mobility UI, (5) aggravation of stress incontinence, and (6) stroke-related incidence of transient incontinence.

The majority of our findings on the risk factors for new-onset FI confirm results of previous studies^11,12^, including increasing stroke severity, advancing age, and increasing pre-morbid disability, similar to those for new onset UI. These risk factors for new-onset FI after stroke are consistent with the major mechanisms of FI after stroke, which include: (1) direct injury to the neural pathways of the brain-gut axis^26–28^ (2) impaired cognition FI, and (3) impaired mobility FI^29^.

As with all clinical research, our study had both strengths and limitations. A notable strength of this study is the consecutive, prospective collection of data, validation of the acute stroke diagnosis by a consultant stroke physician, and the validation of the incontinence diagnosis by a specialist continence nurse. Our study was, however, limited by its single-site location. The study site was a certified Comprehensive Stroke Center (CSC)^30^, and as with other CSCs, may see more patients with more complex conditions. Future multi-site studies may be useful for determining whether the incidence of incontinence after acute stroke varies across the various stroke service levels. Finally, this study is also limited by the absence of an analysis examining the association between stroke comorbidities and incidence of incontinence after stroke. A previous study by Nakayama et al. (1997) on a community-based sample of acute stroke patients identified that hypertension, Diabetes Mellitus (DM), and disabling comorbidities may also be significant predictors of incontinence after stroke^11^.

## CONCLUSION

The current incidence and prevalence rates of urinary and fecal incontinence among adults presenting to an Australian CSC after acute stroke are lower than estimates from the 20^th^ century. Continence assessment, monitoring, and care after acute stroke are more likely to be needed by older patients, those who experienced a severe stroke or intraventricular haemorrhage, and those with greater premorbid disability. Given that incontinence is still relatively common after a stroke, it is important that stroke services continue to provide continence care and rehabilitation.

## Data Availability

Data is available and stored following the ethics approval of this study.

## ACKNOWLEDGEMENTS

None.

## NON-STANDARD ABBREVIATIONS AND ACRONYMS

ANZCTR: Australian New Zealand Clinical Trials Registry
CI: Confidence Interval
CSC: Comprehensive Stroke Centre
CT: Computed Tomography
FI: Fecal Incontinence
ICD-11: International Classification of Diseases eleventh revision
ICS: International Continence Society
MRI: Magnetic Resonance Imaging
OR: Odds Ratio
UI: Urinary Incontinence

